# First-in-Human Study of a First-in-Class AI-Designed Monoclonal Antibody (GB-0669) Against the Conserved SARS-CoV-2 Spike S2 Stem Helix

**DOI:** 10.1101/2025.10.07.25337449

**Authors:** Francesco Borriello, Gavin C.K.W. Koh, Iñaki F. Troconiz, Rounak Nassirpour, Lovely Goyal, Anvar Suyundikov, Akber Safder, Nicholas Robertson, Anna Allen, Andrew Robertson, Stephanie Straley, Pam Farmer, Denise Murphy, Eric Carlin, Kimberly P. Schmitt, Amarendra Pegu, Daria Hazuda, Alexandra Snyder, Dinesh P. De Alwis

## Abstract

**Background:** Antibodies against the SARS-CoV-2 spike receptor-binding domain provided effective COVID-19 treatment until resistant variants emerged. GB-0669 is a half-life extended monoclonal antibody optimized using artificial intelligence, targeting the conserved spike S2 stem helix, a region with limited selective pressure from natural infection-or vaccine-induced antibody responses.

**Methods:** Pre-clinical safety studies were conducted in cynomolgus monkeys. In the first-in-human trial, healthy adults aged 18–55 received single intravenous doses of GB-0669 or placebo in five ascending cohorts (100, 300, 600, 1200, and 2400 mg). Participants were monitored for 43 weeks to evaluate safety, pharmacokinetics (PK), and pharmacodynamics (PD; serum live virus neutralization). *In vitro* studies assessed neutralization of GB-0669 combined with antiviral drugs (remdesivir, nirmatrelvir, and molnupiravir).

**Results:** Pre-clinical studies revealed no safety concerns. In the clinical trial (n=51; 36 GB-0669, 15 placebo), GB-0669 was well-tolerated without dose-limiting toxicities; all adverse reactions were mild (Grade 1 or 2). PK showed dose-proportionality up to 2400 mg, with a half-life of 54 days. Dose-dependent increases in serum live virus neutralization occurred at 600 and 1200 mg, with separation from placebo. The estimated neutralizing index that adjusts GB-0669 serum concentrations for its *in vitro* neutralizing potency supported therapeutic efficacy for two weeks post-administration. Finally, *in vitro* experiments showed improved neutralization profiles of GB-0669 in combination with antivirals.

**Conclusions:** The data support exploring GB-0669 at 1200 mg in a Phase 2 trial for treating COVID-19 in immunocompromised individuals. The combination of GB-0669 with antiviral drugs may offer additional therapeutic benefits.

## Background

During the coronavirus disease 2019 (COVID-19) pandemic, monoclonal antibodies (mAbs) played a key role in the prevention and treatment of severe acute respiratory syndrome coronavirus 2 (SARS-CoV-2) infection, alongside vaccines, small-molecule antivirals, and immunomodulatory drugs [1]. Multiple mAbs were developed for the treatment and prophylaxis of COVID-19 [1–3], but many have since lost effectiveness, and their authorization has been withdrawn [4]. The receptor-binding domain (RBD) of the SARS-CoV-2 spike protein is an immunodominant region and is targeted by the majority of neutralizing antibodies in COVID-19 immune sera [5,6]. Therefore, the RBD is under intense selective evolutionary pressure due to population-level immunity [7], which has led to rapid viral evolution and resistance to all authorized mAbs to-date [4]. Pemivibart is currently the only mAb still authorized for use against COVID-19, however its authorization is limited to pre-exposure prophylaxis rather than treatment. Evidence suggests that, as for other mAbs directed against the RBD, resistance is developing [8].

In the post-pandemic era, immunocompromised patients remain at an increased risk of hospitalization and death, despite high vaccine uptake in this population [9]. The patients at the highest risk of morbidity and death are severely immunocompromised individuals who would not be expected to mount a meaningful antibody response post-vaccination, such as those on B-cell depleting therapy [10,11]. As such, COVID-19 infection is a frequent cause of non-cancer-related mortality in these patients [10,11].

The S2 domain fusion machinery of SARS-CoV-2, comprised of the fusion peptide and stem helix, is not immunodominant and, therefore, not subject to the same selective pressure from the antibody responses elicited by natural infections or vaccines [12]. This allows us to hypothesize that a mAb that binds this domain will have far greater longevity and lower risk of SARS-CoV-2 resistance. Antibodies targeting the S2 stem helix of the spike protein are rare, and exhibit lower neutralization potency than anti-RDB antibodies, thereby constraining their therapeutic potential [13]. To overcome this limitation, we utilized a structure-conditioned machine learning protein design approach integrated with high-throughput experimental assessment of spike binding and pseudovirus neutralization to optimize a mAb directed against the S2 stem-helix of the SARS-CoV-2 spike protein, leading to the identification of the lead candidate GB-0669 [14]. GB-0669 was also designed with an LS mutation in the Fc region, which has been shown to extend an antibody’s half-life and promote translocation to mucosal tissue [15,16]. In keeping with our hypothesis and confirmed by structural and functional analyses, GB-0669 targets a highly conserved epitope and demonstrates robust neutralization activity against all SARS-CoV-2 variants tested to-date and other sarbecoviruses, such as severe acute respiratory syndrome coronavirus 1 (SARS-CoV-1) and Bat SARS-like coronavirus WIV1 [14].

After pre-clinical testing, a healthy volunteer study can examine the safety, tolerability and pharmacokinetics (PK) of an investigative agent. In addition, serum samples can provide surrogate data for *in vivo* activity through *ex vivo* serum neutralizing activity against SARS-CoV-2 [17]. Therefore, it is also possible to evaluate potential anti-viral effectiveness of a therapy.

Here, we report the preclinical safety profile of GB-0669, findings from a healthy-volunteer first-in-human (FIH) study, *ex vivo* serum neutralization activity, and *in vitro* antiviral activity of GB-0669 alone and in combination with small-molecule antivirals. Collectively, these data demonstrate a favorable safety profile and support advancement to a Phase 2 trial to evaluate efficacy for the treatment of COVID-19 in immunocompromised patients.

## Methods

### Pre-clinical toxicity study

Pre-clinical studies were performed in non-human primates by Charles River Laboratories International Inc. (Wilmington, MA, USA) to examine the potential toxicity of GB-0669. Briefly, cynomolgus macaques underwent single or repeated dosing intravenously (IV) of GB-0669 and a comprehensive evaluation of safety measures including clinical, laboratory and pathological assessments. Further details of the procedures are given in the supplementary information.

The study was conducted in accordance with the U.S. Department of Health and Human Services, Food and Drug Administration, United States Code of Federal Regulations, Title 21, Part 58: Good Laboratory Practice for Nonclinical Laboratory Studies and as accepted by Regulatory Authorities throughout the European Union (OECD Principles of Good Laboratory Practice), Japan (MHLW), and other countries that are signatories to the OECD Mutual Acceptance of Data Agreement.

### First-in-human study

#### Study Design and Subjects

This phase 1, randomized, double-blind, placebo-controlled, sequential group, single ascending dose study was undertaken to evaluate the safety, tolerability and PK of GB-0669 in healthy subjects (NCT07050511). The study was conducted at a dedicated phase 1 unit in Orlando, Florida between July 2023 and November 2024. Fifty-one healthy adult volunteers were enrolled and randomized to receive either single dose GB-0669 or placebo IV in five sequential ascending cohorts of 100 mg, 300 mg, 600 mg, 1200 mg, and 2400 mg in ratios of 3:3, 3:3, 10:3, 10:3, 10:3 respectively. Participants were followed for 43 weeks (up to and including Day 302; a minimum of five-half-lives estimated from the available pre-clinical data).

Healthy male or female participants were eligible if aged 18–55 years at screening, without clinically significant abnormalities (as determined by the Investigator) at screening and pre-dose on Day 1, and negative SARS-CoV-2 rapid antigen or polymerase chain reaction (PCR) tests prior to randomization. Written informed consent was required from all participants before any study-related activities were carried out. The study followed the Declaration of Helsinki ethical guidelines and the International Council for Harmonisation Guidelines for Good Clinical Practice. The full inclusion and exclusion criteria are available in the supplementary material.

#### Safety

The safety of all participants enrolled in this trial was carefully monitored. The investigator(s), medical monitor, the Sponsor, and an independent safety review committee (iSRC) reviewed participants’ safety assessments, adverse events (AEs), laboratory tests, and monitored for any dose limiting toxicities (DLTs). Further details of the safety analysis are provided in the Supplementary information.

#### Pharmacokinetics

A validated electrochemiluminescence (ECL) assay quantitated GB-0669 in human serum. The immunoassay employed MSD Gold Streptavidin plates (Miso Scale Discovery, Rockville, MD) coated with GB-0669 anti-idiotypic antibody as the capture reagent. Resulting ECL units were fitted using a 5-parameter logistic (5-PL) model with 1/y^2^ weighting to calculate the GB-0669 concentrations in both the quality control and study samples. The calibration curve range of this method is 15000 ng/mL to 100 ng/mL in 100% pooled human serum. The PK Set consists of all participants treated with any dose of study drug.

#### Anti-Drug Antibody Assays

Serum samples for the detection of anti-drug antibodies (ADAs) were processed by using a 3-tier testing scheme using a validated ECL solution-phase bridging method. Diluted samples were incubated in a solution phase with biotinylated GB-0669 and ruthenium ([sulfo-TAG]-labeled GB-0669). Samples were reported as positive for ADA in the screening assay if the mean ECL value was at or above the ECL value of the plate-specific cut point factor. Screen-positive samples were retested in a confirmation assay, where samples were analyzed in the presence of excess drug to determine if the sample’s positive response was specific to GB-0669.

#### Live Virus Neutralization

SARS-CoV-2 *ex vivo* neutralizing activity in serum samples was determined using a validated proprietary microneutralization assay (Cerba Research Netherlands, Rotterdam, the Netherlands). Triplicate serial dilutions of serum samples were mixed with a fixed amount of a reference SARS-CoV-2 virus strain (BetaCoV/Germany/BavPat1/2020, BA5.5 strain hCoV-19/USA/COR-22-063113/2022, or XBB1.5 strain hCoV-19/USA/MD-HP40900/2022) and incubated for 1 hour at 37°C. The mixture was then added to 96-well microtiter plates containing VeroE6 cells at sub-confluent density and again incubated for 1 hour at 37°C. The inoculum mixture was then replaced by infection medium and incubation continued for 16-24 hours at 37°C in the presence of a carboxymethylcellulose overlay. Cells subsequently underwent formalin fixation and viral microplaques were detected via immunostaining with an anti-nucleoprotein antibody, a peroxidase conjugate, and TrueBlue staining. Microplaques were imaged and counted in an SX Ultimate-V Analyzer (Cellular Technology Limited, Shaker Heights, USA). Neutralization titers were calculated from these data according to the method described by Zielinska et al. [18]. This method uses the sample dilutions directly above and below the reduction point (50%) and the spot counts corresponding with those dilutions are used to calculate the microneutralization (MN50) titer.

The neutralizing activity of GB-0669, at a starting final concentration of 18 μg/mL, was tested *in vitro* using SARS-CoV-2 Omicron JN.1 (strain SARS-CoV-2 hCoV-19/USA/NY/PV96109/2023 (JN.1) (Omicron JN.1)) at Virology Research Services Ltd (Sittingbourne, United Kingdom).

Remdesivir (Selleck Chemicals, TX, USA; S8932), nirmatrelvir (Insight Biotechnology Limited, United Kingdom, HY-138687), and molnupiravir (EIDD-1931; Insight Biotechnology Limited, HY-125033) was assessed at a starting final concentration of 20 μM, both individually and in combination with GB-0669. Neutralization was measured using 12-point titration curves in technical triplicate with 3-fold dilutions on Vero cells. Following 72 hours incubation, an MTT assay (Invitrogen, M6494) was used to assess cell viability using a Promega GloMax Explorer System plate reader at 560 nm. The percentage inhibition of SARS-CoV-2 Omicron JN.1 infection was calculated using the following formula:

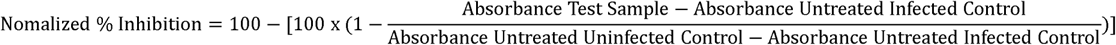

Half maximal effective concentration (EC50) values were derived for each Log-transformed antibody titration curve using log(inhibitor) vs. response -- Variable slope (four parameters) equation using GraphPad Prism software (v10).

### Statistical analysis

#### Safety

Quantitative safety data and frequency counts were compiled for classification of qualitative safety data. Safety data were summarized by treatment arms with placebo pooled from cohorts.

#### Population pharmacokinetic analysis

The analysis of the time course of the serum concentrations of GB-0669 was performed using NONMEM software (version 7.5; Icon Development Solutions, Hanover, MD, USA) with the First Order Conditional Estimation (FOCE) method and the INTERACTION option. Given the homogenous subject population, no covariate analysis was attempted.

Serum levels were logarithmically transformed for analysis. Interindividual variability (IIV) was modelled using the exponential model and the significance of the non-diagonal elements of the variance-covariance matrix was evaluated. The residual error was characterized by an additive model on the logarithmic scale.

Model selection was guided by the minimum value of the objective function, parameter precision, and visual inspection of the goodness of fit plots. A reduction of 3.84 and 6.61 points in the minimum value of the objective function (approximately equal to-2×Log[likelihood[[-2LL])] between two hierarchical models differing in one parameter was considered significant at the 5% and 1% levels of significance, respectively. Precision of model parameters was evaluated calculating the coefficient of variation as the ratio between the standard error and the estimate of the parameter and multiplied by 100.

The pharmacokinetic disposition of GB-0669 in serum was described with compartmental models parameterized in terms of apparent volumes of distribution, distribution (inter-compartmental) clearance, and total elimination clearance. The selected models were evaluated using a simulation-based diagnostic generated from simulated serum GB-0669 concentrations for one thousand patients for each dose group. At each timepoint, the median and the 90% prediction interval were calculated and presented graphically, together with the raw data.

#### Neutralizing index analysis

The potency of GB-0669 against SARS-CoV-2 variants, as a measure of its predicted efficacy to protect patients with symptomatic COVID-19 progressing to hospitalization, was calculated as a neutralizing index based on a modified calculation by Stadler et al. [19]. The selected pharmacokinetic model was used to investigate the exposure relative to *in vitro* efficacy-related parameters such as EC50 or neutralizing index. The latter was calculated using the following expression:

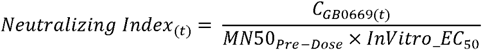

MN50_Pre-dose_ is the median value of the MN50 titers calculated using pre-dose observations for each of the Omicron variants and equal to 94 and 114.5 for XBB.1.5 and BA.5.5, respectively. C_GB-0669(t)_ are the simulated serum concentrations of GB-0669 over time obtained from one thousand virtual subjects receiving a short IV infusion of 1200 mg. The *in vitro* EC50 values for GB-0669 were determined by evaluating its neutralization activity against live SARS-CoV-2 BA.5 and XBB.1.1 variants using an immunofluorescence-based microneutralization assay as previously described [14]. The resulting EC50 values were 0.048 μg/mL for BA.5 and 0.053 μg/mL for XBB.1.1. This calculation used the predicted plasma concentration over a course of two weeks at 1200 mg.

## Results

### Pre-clinical toxicology

IV administration of GB-0669 was well-tolerated in cynomolgus macaques both as a single-or repeat-dose administration. A single IV dose of 400 mg/kg GB-0669 was associated with very slight edema (Grade 1) in males and well-defined erythema (Grade 2) in both sexes at the injection site in the majority of GB-0669-treated animals. There were no adverse findings, and the no-observed-adverse-effect-level (NOAEL) was determined to be 400/600 mg/kg (the doses given in the repeated dose schedule) which corresponds to Day 23 combined-sex, peak and cumulative systemic exposures of 18,700,000 ng/mL, and 1,610,000,000 ng.hr/mL [peak serum concentration (C_max_) and area under the curve (AUC)_0–528hr_, respectively].

Systemic exposure to GB-0669 appeared to be independent of sex for the single dose and repeated dose groups. Following a single IV infusion of GB-0669 on Day 1, mean C_max_, AUC_0–1416hr_, and AUC_0-inf_ values were 10,000,000 ng/mL, 1,890,000,000 hr.ng/mL, and 1,970,000,000 hr.ng/mL, respectively. In the repeat dose group, following the first IV infusion of GB-0669 on Day 1, mean C_max_, and AUC_0-528hr_ values were 10,200,000 ng/mL, and 1,170,000,000 hr.ng/mL, respectively. Following the second IV infusion of GB-0669 on Day 23, mean C_max_, AUC_0-528hr_, and AUC_0-2184hr_ values were 18,700,000 ng/mL, 1,610,000,000 hr.ng/mL, and 2,290,000,000 hr.ng/mL, respectively. Systemic exposure (based on dose-normalized C_max_ and AUC_0-528hr_ values) to GB-0669 did not appear to change following repeated GB-0669 administration.

Moreover, no anti-GB-0669 antibodies were detected in GB-0669-treated animals.

### First-in-human study

Following the favorable pre-clinical toxicity data, 155 subjects were considered for inclusion and finally 51 participants were enrolled in the FIH study with a median age of 36 years (range 23 to 55 years) as shown in **Supplementary Figure 1**. Detailed demographics and baseline characteristics are shown in **Supplementary Table 1**.

#### Safety

**Table 1** details the occurrence of all treatment-emergent adverse events (TEAEs), both those that were considered treatment-related and those considered unrelated to treatment in both placebo and treatment groups. No dose-limiting toxicities were observed in any group and all treatment-related TEAEs observed were mild (Grade 1 or 2). One Grade 2 AE of an infusion related reaction (IRR) in the 1200 mg GB-0669 cohort was reported, and the infusion was interrupted. Four combined Grade 3 and 4 AEs of elevated creatine phosphokinase associated with physical exertion were observed. No serious AEs (SAEs) were observed.

**Table 1.**
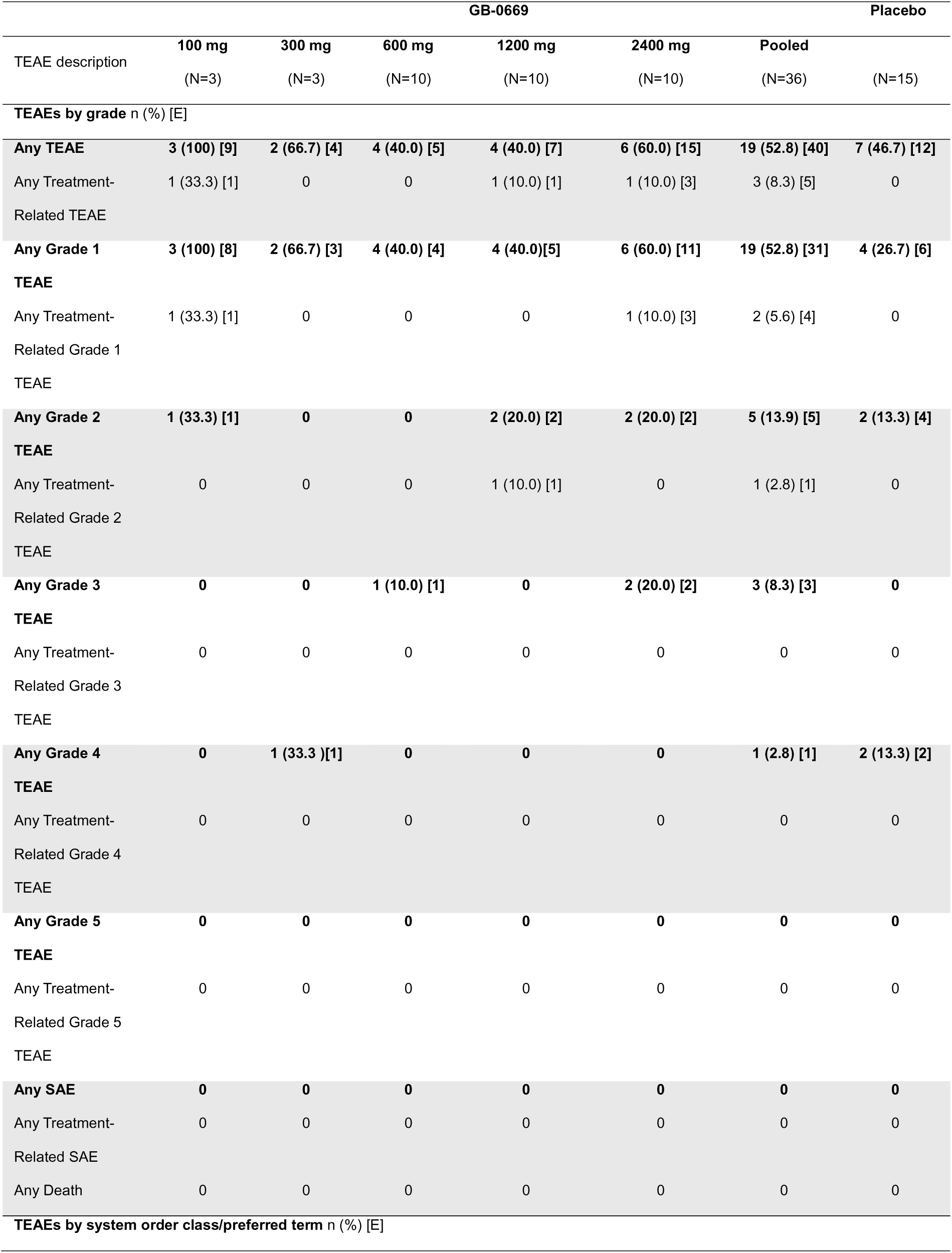
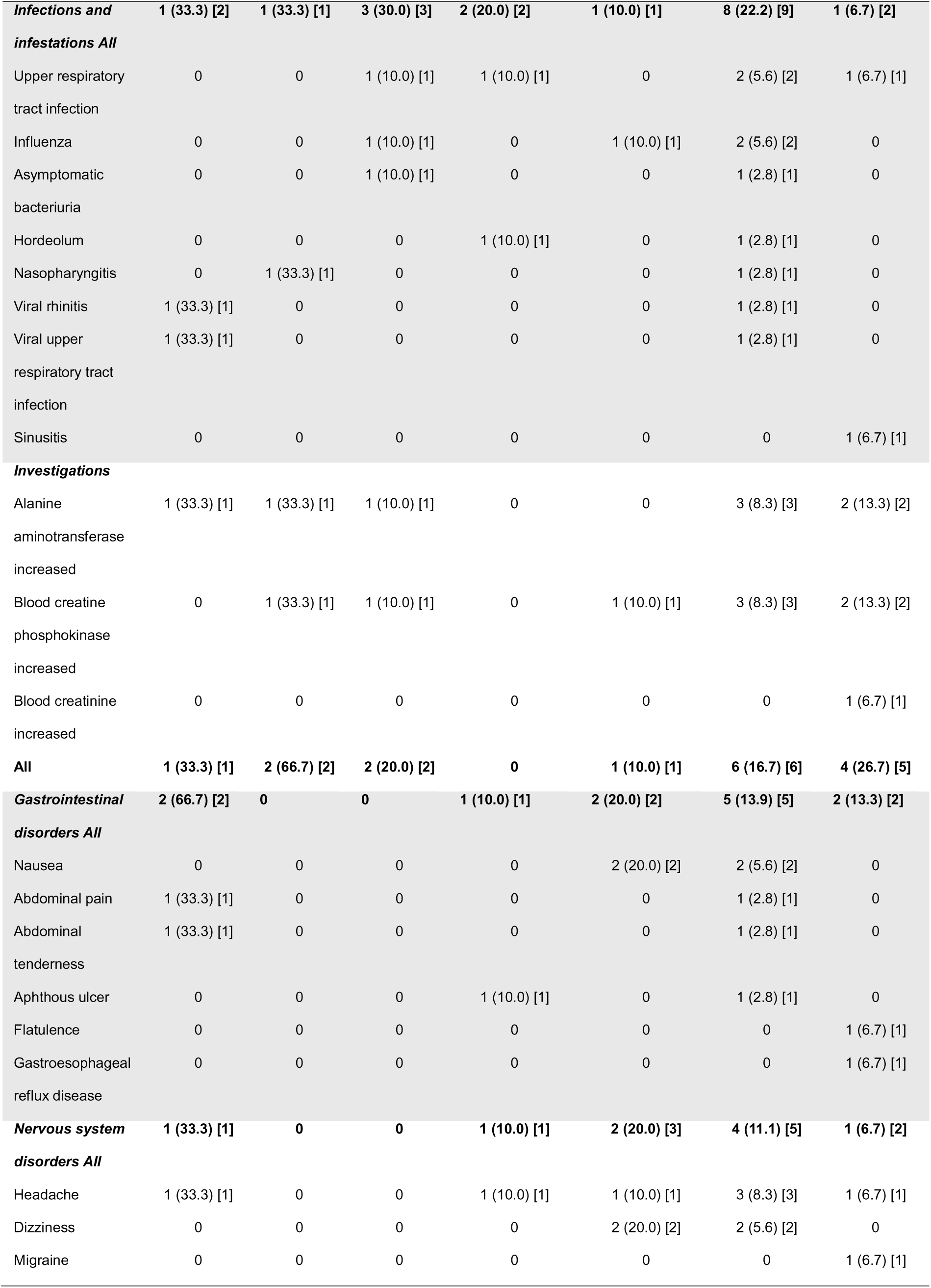
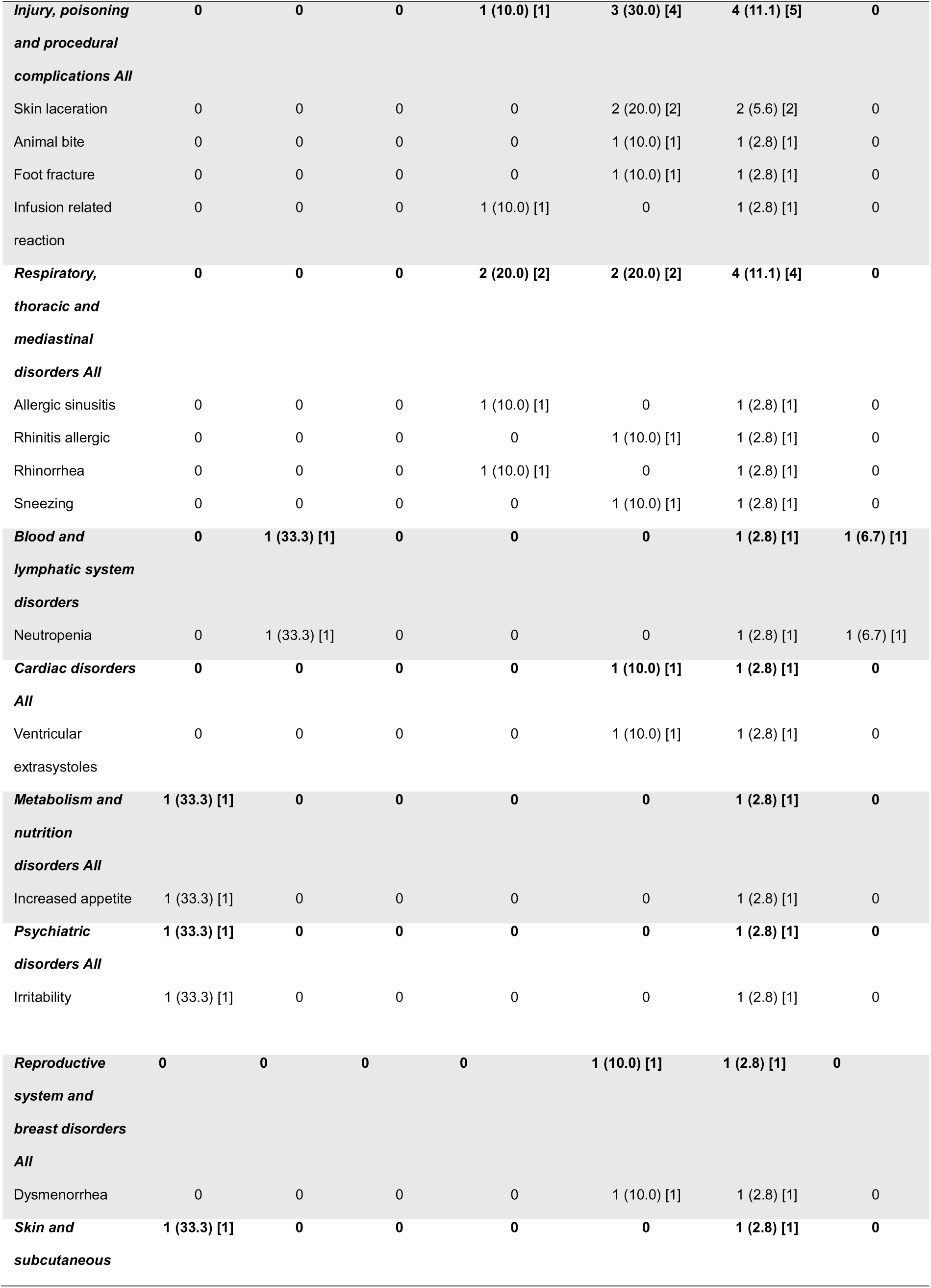
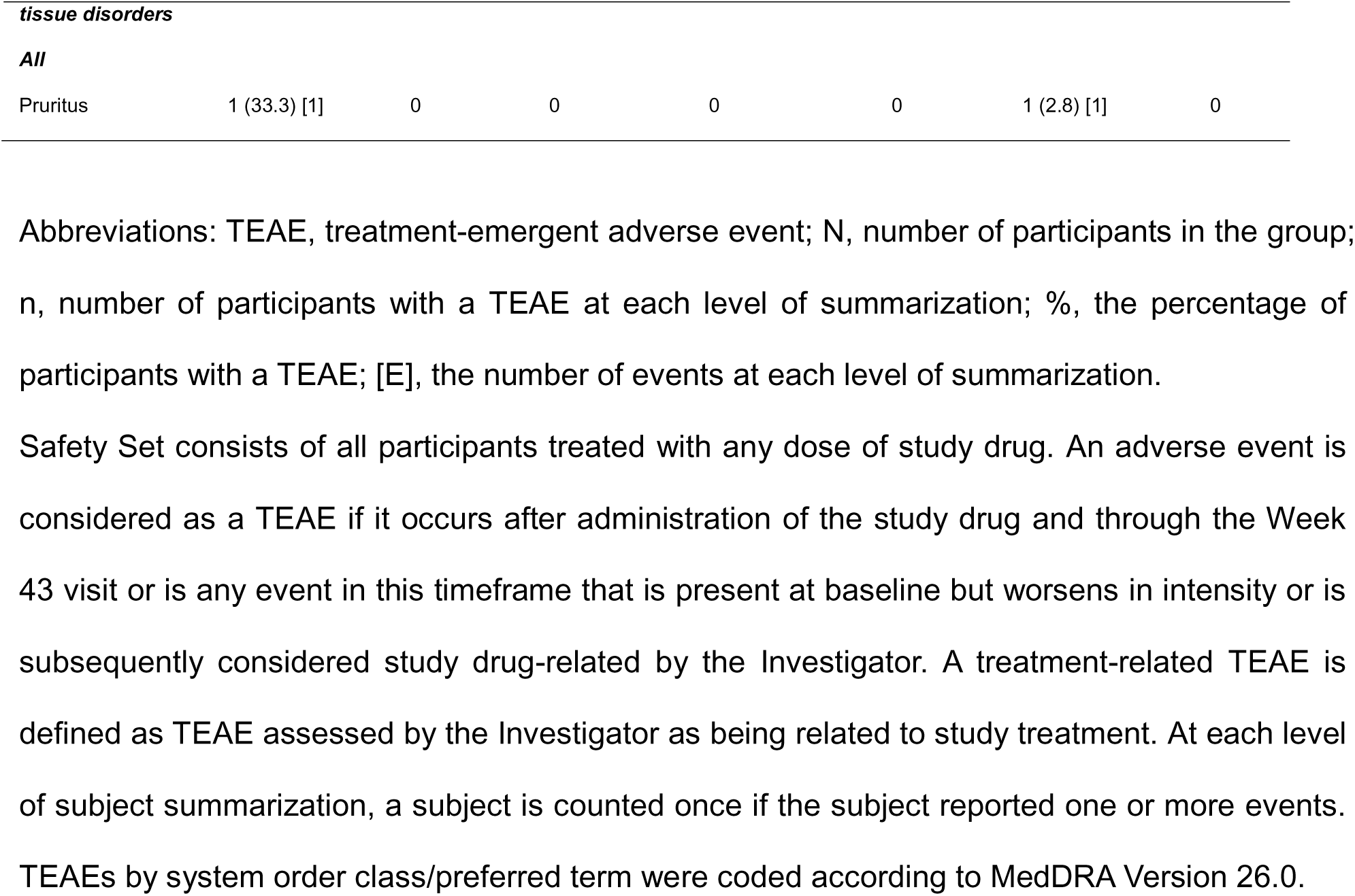
Summary of treatment-emergent adverse events.

|  | GB-0669 |  |  |  |  | Placebo |  |
| --- | --- | --- | --- | --- | --- | --- | --- |
| TEAE description | 100 mg<br>(N=3) | 300 mg<br>(N=3) | 600 mg<br>(N=10) | 1200 mg<br>(N=10) | 2400 mg<br>(N=10) | Pooled<br>(N=36) | (N=15) |
| **TEAEs by grade n (%) [E]** |  |  |  |  |  |  |  |
| **Any TEAE** | **3 (100) [9]** | **2 (66.7) [4]** | **4 (40.0) [5]** | **4 (40.0) [7]** | **6 (60.0) [15]** | **19 (52.8) [40]** | **7 (46.7) [12]** |
| Any Treatment-Related TEAE | 1 (33.3) [1] | 0 | 0 | 1 (10.0) [1] | 1 (10.0) [3] | 3 (8.3) [5] | 0 |
| **Any Grade 1**<br>**TEAE** | **3 (100) [8]** | **2 (66.7) [3]** | **4 (40.0) [4]** | **4 (40.0) [5]** | **6 (60.0) [11]** | **19 (52.8) [31]** | **4 (26.7) [6]** |
| Any Treatment-Related Grade 1<br>TEAE | 1 (33.3) [1] | 0 | 0 | 0 | 1 (10.0) [3] | 2 (5.6) [4] | 0 |
| **Any Grade 2**<br>**TEAE** | **1 (33.3) [1]** | **0** | **0** | **2 (20.0) [2]** | **2 (20.0) [2]** | **5 (13.9) [5]** | **2 (13.3) [4]** |
| Any Treatment-Related Grade 2<br>TEAE | 0 | 0 | 0 | 1 (10.0) [1] | 0 | 1 (2.8) [1] | 0 |
| **Any Grade 3**<br>**TEAE** | **0** | **0** | **1 (10.0) [1]** | **0** | **2 (20.0) [2]** | **3 (8.3) [3]** | **0** |
| Any Treatment-Related Grade 3<br>TEAE | 0 | 0 | 0 | 0 | 0 | 0 | 0 |
| **Any Grade 4**<br>**TEAE** | **0** | **1 (33.3) [1]** | **0** | **0** | **0** | **1 (2.8) [1]** | **2 (13.3) [2]** |
| Any Treatment-Related Grade 4<br>TEAE | 0 | 0 | 0 | 0 | 0 | 0 | 0 |
| **Any Grade 5**<br>**TEAE** | **0** | **0** | **0** | **0** | **0** | **0** | **0** |
| Any Treatment-Related Grade 5<br>TEAE | 0 | 0 | 0 | 0 | 0 | 0 | 0 |
| **Any SAE** | **0** | **0** | **0** | **0** | **0** | **0** | **0** |
| Any Treatment-Related SAE | 0 | 0 | 0 | 0 | 0 | 0 | 0 |
| Any Death | 0 | 0 | 0 | 0 | 0 | 0 | 0 |
| **TEAEs by system order class/preferred term n (%) [E]** |  |  |  |  |  |  |  |
| **Infections and infestations All** | **1 (33.3) [2]** | **1 (33.3) [1]** | **3 (30.0) [3]** | **2 (20.0) [2]** | **1 (10.0) [1]** | **8 (22.2) [9]** | **1 (6.7) [2]** |
| Upper respiratory tract infection | 0 | 0 | 1 (10.0) [1] | 1 (10.0) [1] | 0 | 2 (5.6) [2] | 1 (6.7) [1] |
| Influenza | 0 | 0 | 1 (10.0) [1] | 0 | 1 (10.0) [1] | 2 (5.6) [2] | 0 |
| Asymptomatic bacteriuria | 0 | 0 | 1 (10.0) [1] | 0 | 0 | 1 (2.8) [1] | 0 |
| Hordeolum | 0 | 0 | 0 | 1 (10.0) [1] | 0 | 1 (2.8) [1] | 0 |
| Nasopharyngitis | 0 | 1 (33.3) [1] | 0 | 0 | 0 | 1 (2.8) [1] | 0 |
| Viral rhinitis | 1 (33.3) [1] | 0 | 0 | 0 | 0 | 1 (2.8) [1] | 0 |
| Viral upper respiratory tract infection | 1 (33.3) [1] | 0 | 0 | 0 | 0 | 1 (2.8) [1] | 0 |
| Sinusitis | 0 | 0 | 0 | 0 | 0 | 0 | 1 (6.7) [1] |
| **Investigations** |  |  |  |  |  |  |  |
| Alanine aminotransferase increased | 1 (33.3) [1] | 1 (33.3) [1] | 1 (10.0) [1] | 0 | 0 | 3 (8.3) [3] | 2 (13.3) [2] |
| Blood creatine phosphokinase increased | 0 | 1 (33.3) [1] | 1 (10.0) [1] | 0 | 1 (10.0) [1] | 3 (8.3) [3] | 2 (13.3) [2] |
| Blood creatinine increased | 0 | 0 | 0 | 0 | 0 | 0 | 1 (6.7) [1] |
| **All** | **1 (33.3) [1]** | **2 (66.7) [2]** | **2 (20.0) [2]** | **0** | **1 (10.0) [1]** | **6 (16.7) [6]** | **4 (26.7) [5]** |
| **Gastrointestinal disorders All** | **2 (66.7) [2]** | **0** | **0** | **1 (10.0) [1]** | **2 (20.0) [2]** | **5 (13.9) [5]** | **2 (13.3) [2]** |
| Nausea | 0 | 0 | 0 | 0 | 2 (20.0) [2] | 2 (5.6) [2] | 0 |
| Abdominal pain | 1 (33.3) [1] | 0 | 0 | 0 | 0 | 1 (2.8) [1] | 0 |
| Abdominal tenderness | 1 (33.3) [1] | 0 | 0 | 0 | 0 | 1 (2.8) [1] | 0 |
| Aphthous ulcer | 0 | 0 | 0 | 1 (10.0) [1] | 0 | 1 (2.8) [1] | 0 |
| Flatulence | 0 | 0 | 0 | 0 | 0 | 0 | 1 (6.7) [1] |
| Gastroesophageal reflux disease | 0 | 0 | 0 | 0 | 0 | 0 | 1 (6.7) [1] |
| **Nervous system disorders All** | **1 (33.3) [1]** | **0** | **0** | **1 (10.0) [1]** | **2 (20.0) [3]** | **4 (11.1) [5]** | **1 (6.7) [2]** |
| Headache | 1 (33.3) [1] | 0 | 0 | 1 (10.0) [1] | 1 (10.0) [1] | 3 (8.3) [3] | 1 (6.7) [1] |
| Dizziness | 0 | 0 | 0 | 0 | 2 (20.0) [2] | 2 (5.6) [2] | 0 |
| Migraine | 0 | 0 | 0 | 0 | 0 | 0 | 1 (6.7) [1] |
| ***Injury, poisoning and procedural complications All*** | **0** | **0** | **0** | **1 (10.0) [1]** | **3 (30.0) [4]** | **4 (11.1) [5]** | **0** |
| Skin laceration | 0 | 0 | 0 | 0 | 2 (20.0) [2] | 2 (5.6) [2] | 0 |
| Animal bite | 0 | 0 | 0 | 0 | 1 (10.0) [1] | 1 (2.8) [1] | 0 |
| Foot fracture | 0 | 0 | 0 | 0 | 1 (10.0) [1] | 1 (2.8) [1] | 0 |
| Infusion related reaction | 0 | 0 | 0 | 1 (10.0) [1] | 0 | 1 (2.8) [1] | 0 |
| ***Respiratory, thoracic and mediastinal disorders All*** | **0** | **0** | **0** | **2 (20.0) [2]** | **2 (20.0) [2]** | **4 (11.1) [4]** | **0** |
| Allergic sinusitis | 0 | 0 | 0 | 1 (10.0) [1] | 0 | 1 (2.8) [1] | 0 |
| Rhinitis allergic | 0 | 0 | 0 | 0 | 1 (10.0) [1] | 1 (2.8) [1] | 0 |
| Rhinorrhea | 0 | 0 | 0 | 1 (10.0) [1] | 0 | 1 (2.8) [1] | 0 |
| Sneezing | 0 | 0 | 0 | 0 | 1 (10.0) [1] | 1 (2.8) [1] | 0 |
| ***Blood and lymphatic system disorders*** | **0** | **1 (33.3) [1]** | **0** | **0** | **0** | **1 (2.8) [1]** | **1 (6.7) [1]** |
| Neutropenia | 0 | 1 (33.3) [1] | 0 | 0 | 0 | 1 (2.8) [1] | 1 (6.7) [1] |
| ***Cardiac disorders All*** | **0** | **0** | **0** | **0** | **1 (10.0) [1]** | **1 (2.8) [1]** | **0** |
| Ventricular extrasystoles | 0 | 0 | 0 | 0 | 1 (10.0) [1] | 1 (2.8) [1] | 0 |
| ***Metabolism and nutrition disorders All*** | **1 (33.3) [1]** | **0** | **0** | **0** | **0** | **1 (2.8) [1]** | **0** |
| Increased appetite | 1 (33.3) [1] | 0 | 0 | 0 | 0 | 1 (2.8) [1] | 0 |
| ***Psychiatric disorders All*** | **1 (33.3) [1]** | **0** | **0** | **0** | **0** | **1 (2.8) [1]** | **0** |
| Irritability | 1 (33.3) [1] | 0 | 0 | 0 | 0 | 1 (2.8) [1] | 0 |
| ***Reproductive system and breast disorders All*** | **0** | **0** | **0** | **0** | **1 (10.0) [1]** | **1 (2.8) [1]** | **0** |
| Dysmenorrhea | 0 | 0 | 0 | 0 | 1 (10.0) [1] | 1 (2.8) [1] | 0 |
| ***Skin and subcutaneous*** | **1 (33.3) [1]** | **0** | **0** | **0** | **0** | **1 (2.8) [1]** | **0** |
| Pruritus | 1 (33.3) [1] | 0 | 0 | 0 | 0 | 1 (2.8) [1] | 0 |

Abbreviations: TEAE, treatment-emergent adverse event; N, number of participants in the group; n, number of participants with a TEAE at each level of summarization; %, the percentage of participants with a TEAE; [E], the number of events at each level of summarization.
Safety Set consists of all participants treated with any dose of study drug. An adverse event is considered as a TEAE if it occurs after administration of the study drug and through the Week 43 visit or is any event in this timeframe that is present at baseline but worsens in intensity or is subsequently considered study drug-related by the Investigator. A treatment-related TEAE is defined as TEAE assessed by the Investigator as being related to study treatment. At each level of subject summarization, a subject is counted once if the subject reported one or more events. TEAEs by system order class/preferred term were coded according to MedDRA Version 26.0.

#### Pharmacokinetics

Following IV infusion, GB-0669 pharmacokinetic data showed two compartment behavior with a rapid distribution phase and a long terminal elimination phase of approximately 54 days (**Figure 1**). **Table 2** lists the population model parameter estimates. All parameters, fixed and random effect, were obtained with high precision as shown by the relative standard errors <25%.

**Figure 1.**
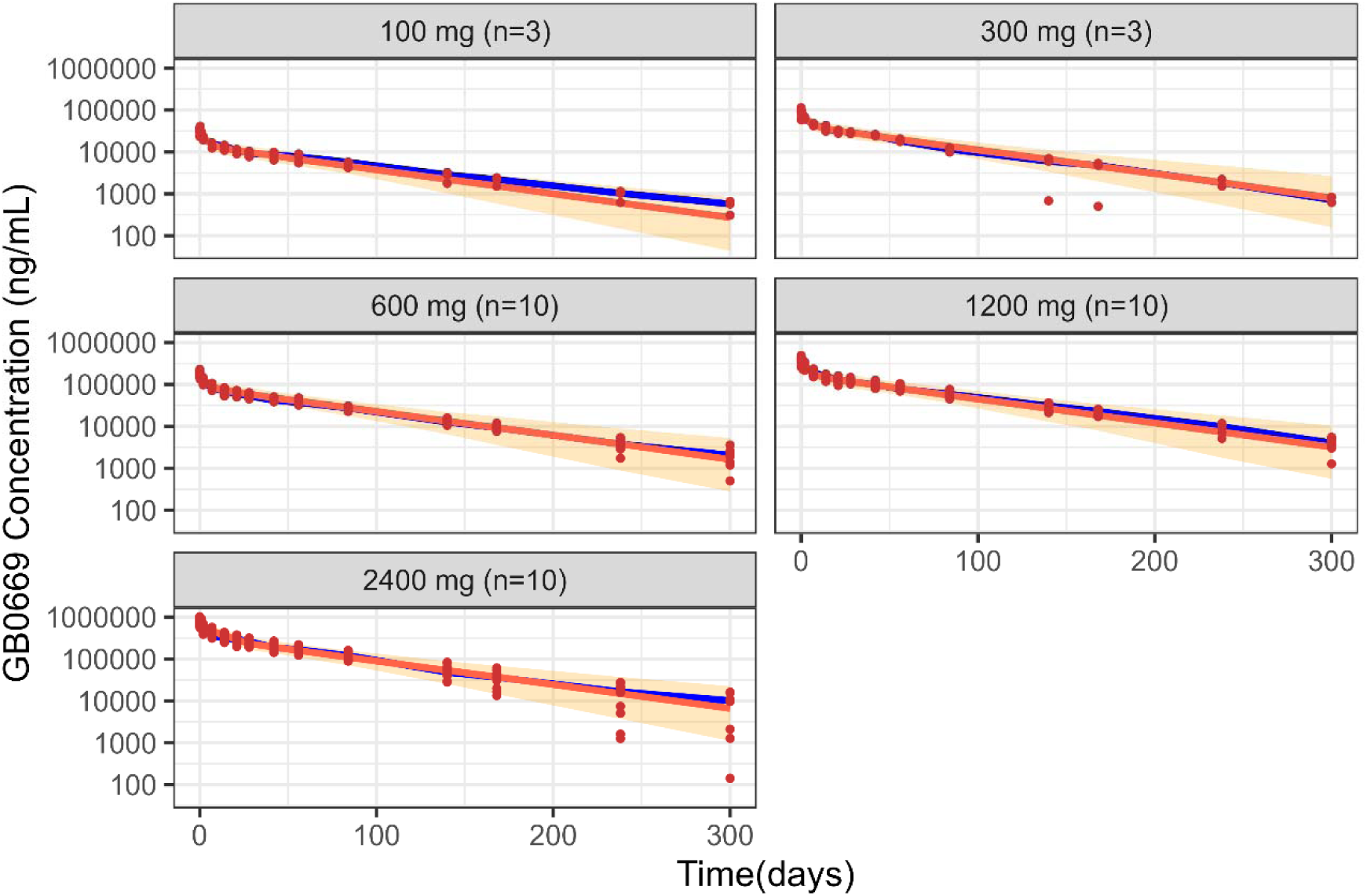
Pharmacokinetic analysis of GB-0669 in healthy participants. Results from simulation-based diagnostics where 1000 virtual profiles were simulated for each dose level. Solid circles represent observed concentrations; Solid lines in blue and red correspond to the median of the observed and simulated profiles, respectively; Shaded areas cover the 90% prediction intervals; n, number of subjects per dose group.

**Table 2.** Population pharmacokinetic model parameters of GB-0669 across all doses.

| Parameter | Typical estimates | IIV (%) | Shrinkage (%) |
| --- | --- | --- | --- |
| CL (L/d) | 0.088 (2.9) | 16.8 (12.1) | 27 |
| V<sub>1</sub> (L) | 3.42 (2) | 10.6 (15.8) | 16 |
| V<sub>2</sub> (L) | 3.1 (7.9) | 37.7 (22) | 6 |
| CL<sub>D</sub> (L/d) | 0.517 (8.2) | - | - |
| Error [log(ng/mL)] | 0.143 (15.2) | - | 7.6 |

Abbreviations: IIV, inter-individual variability; CL, total serum elimination clearance; V<sub>1</sub>, apparent
volume of distribution of the central compartment; V<sub>2</sub>, apparent volume of distribution of the
peripheral compartment; CL<sub>D</sub>, inter-compartment (distribution) clearance between the central
and peripheral compartments.
Values within brackets show the relative standard error (%); IIV is expressed as coefficient of
variation (CV, %) calculated as $\sqrt{e^{\omega^2} - 1} \times 100$ where $\omega^2$ corresponds to the variance of the
random effects; Error is equivalent to a CV (%) of 5%.

The total apparent volume of distribution (V_1_ + V_2_ = 6.34 L), where V_1_ and V_2_ are respectively the apparent volumes of distribution of the central and peripheral compartments, was lower than total body water volume in the adult healthy subject population. The typical value of the terminal half-life derived from the parameter estimates was 53.5 days and ranging from 24 to 65 days in the studied subject population. Estimates of IIV were low for V_1_ and the total elimination serum clearance, CL, (<20%) and moderate for V_2_ (40% approximately). The magnitude of the residual error listed in **Table 2** corresponds to a coefficient of variation of 1.3%. The results of the simulation-based diagnostics summarized in **Figure 1** indicated that the selected model described both the median tendency of the data and their dispersion at any dose level, without model misspecification indicating clear dose proportionality.

#### Anti-drug antibodies

The incidence of treatment-emergent ADAs following GB-0669 administration was found to be 11% (4 of 36 participants treated with GB-0669) over the entire dose range (100mg – 2400 mg).

This compares to 20% in the placebo group (3 of 15 participants treated with placebo). There was no impact on PK as depicted in **Figure 1**.

#### Pharmacodynamic

An exploratory analysis of serum live virus neutralization is presented in **Figure 2** which shows the fold increase in MN50 from baseline against the XBB.1.5 and BA.5.5 Omicron variants. GB-0669 demonstrated a clear dose-response relationship, with meaningful separation from placebo observed at doses of 600 mg and 1200 mg. A single infusion of 1200 mg GB-0669 achieved sustained neutralizing activity, maintaining approximately 10-fold and 5-fold increases from baseline for XBB.1.5 and BA.5.5 variants, respectively, for over two weeks in participants with detectable pre-dose titers.

**Figure 2.**
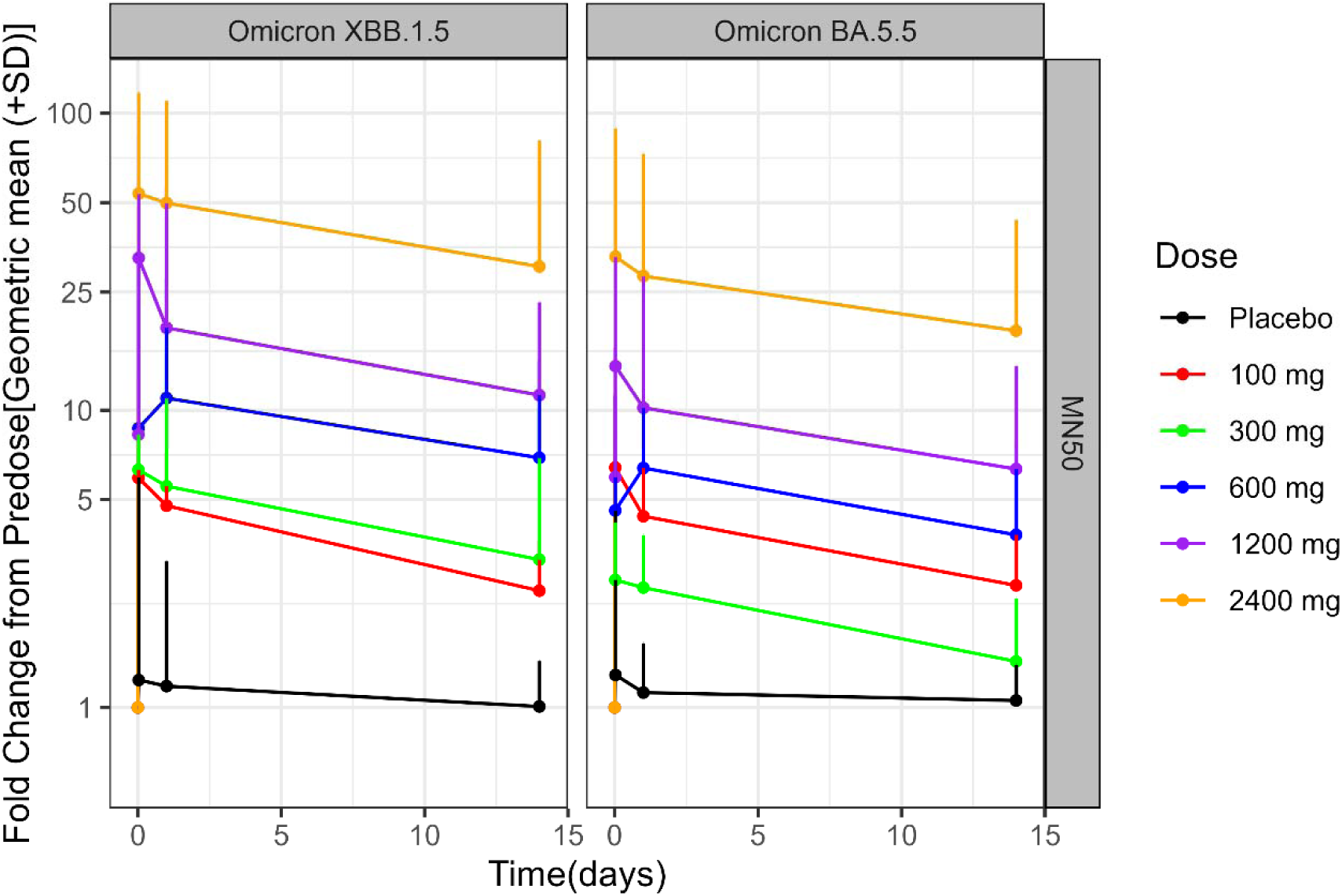
Pharmacodynamic analysis of GB-0669. Analysis of GB-0669 serum live virus neutralization against SARS-CoV-2 Omicron XBB.1.5 and BA.5.5 variants. The graphs show the fold change of the MN50 geometric mean from Predose over time. Placebo is indicated in black with red for 100 mg, green for 300 mg, blue for 600 mg, purple for 1200 mg, and orange for 2400 mg GB-0669 doses. Bars show standard deviation (SD).

#### Neutralizing Index

The neutralizing index was estimated to range from over 75 to above 15 over the course of the 2-week post-infusion phase (**Figure 3**) including 90% prediction intervals. This suggests the activity conferred by GB-0669 remains well above the desired therapeutic threshold of 1 (based on efficacy data from multiple mAbs and convalescent plasma for the treatment period for the overall population) [19]. Collectively, these results support the potential efficacy of GB-0669 as a treatment for COVID-19.

**Figure 3.**
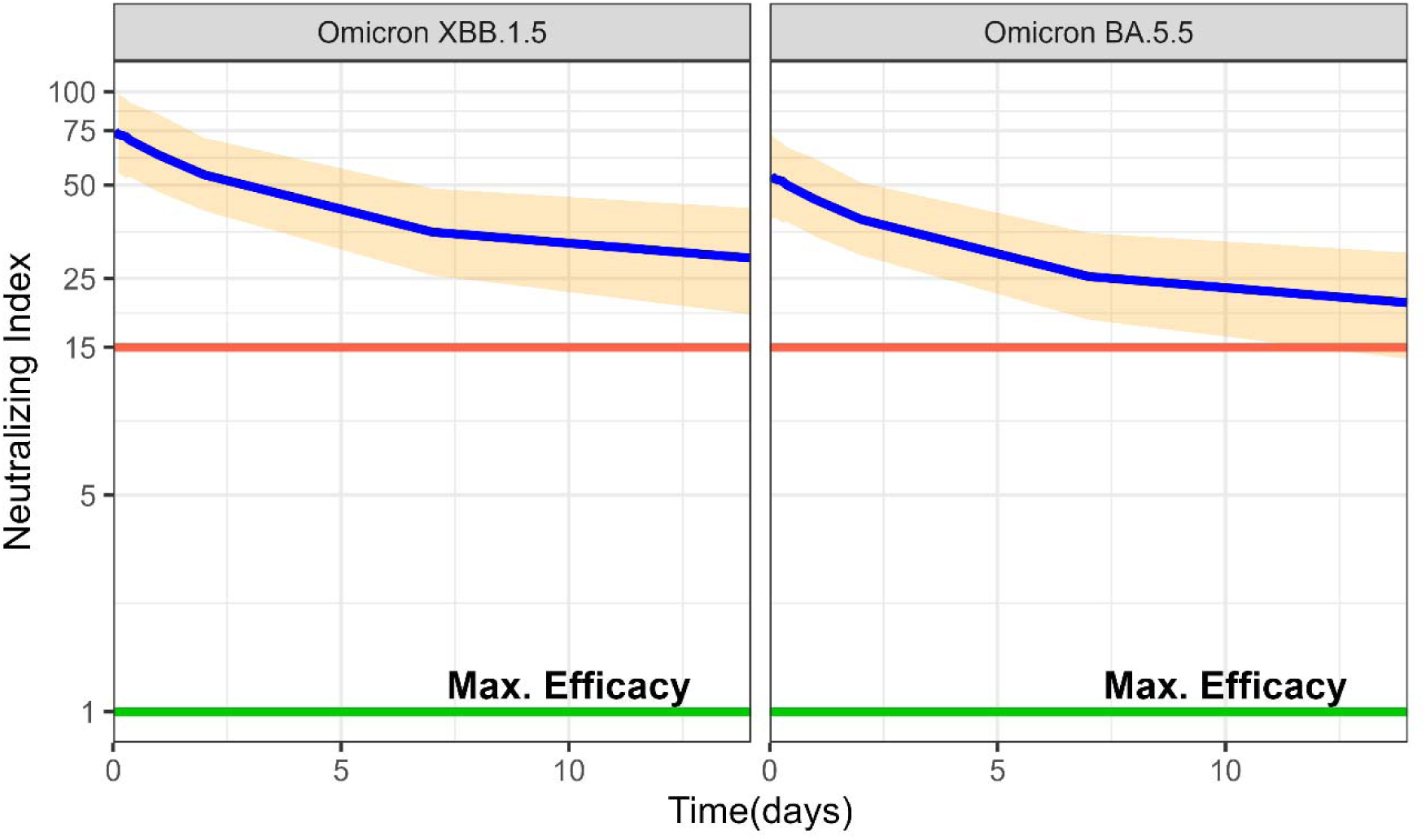
Neutralizing index of GB-0669 against SARS-CoV-2 Omicron XBB.1.5 and BA.5.5 variants which remains above 15 over the course of 2 weeks. Shaded areas cover the neutralizing index 90% prediction intervals corresponding to the Omicron XBB.1.5 and BA.5.5 variants, obtained from 1000 simulated pharmacokinetic profiles assuming a 45 min intravenous infusion of 1200 mg of GB-0669. Solid lines in blue represent the median neutralizing index profiles over a 2-week period; solid lines in green represent maximum therapeutic threshold.

#### In vitro neutralization of GB-0669 in Combination with Antivirals

Small-molecule antivirals, including remdesivir, nirmatrelvir, and molnupiravir, are authorized for COVID-19 treatment. Pre-clinical studies and emerging clinical observations suggest that combining monoclonal antibodies with antivirals can enhance viral neutralization and potentially improve clinical outcomes, particularly in immunocompromised patients [20]. However, human data thus far have primarily consisted of anecdotal case reports or small cohort studies, rather than large, randomized trials. To build on these observations, we evaluated the neutralization activity of GB-0669 combined with remdesivir, nirmatrelvir, or molnupiravir against the SARS-CoV-2 Omicron JN.1 variant. These combinations demonstrated superior neutralization profiles, with improved potency and efficacy compared to either GB-0669 or antivirals alone (**Table 3**).

**Table 3.** EC50 values for neutralization of SARS-CoV-2 JN.1 by GB-0669 as a single agent and in combination with Remdesivir, Nirmatrelvir, and Molnupiravir.

| Condition | Remdesivir |  | Nirmatrelvir |  | Molnupiravir |  |
| --- | --- | --- | --- | --- | --- | --- |
|  | EC50<br>(µg/mL) | Upper plateau<br>(% inhibition)<sup>c</sup> | EC50<br>(µg/mL) | Upper plateau<br>(% inhibition)<sup>c</sup> | EC50<br>(µg/mL) | Upper plateau<br>(% inhibition)<sup>c</sup> |
| **GB-0669 \| Single agent** | 0.077 | 104.9 | 0.089 | 89.14 | 0.69 | 115.6 |
| **Antiviral \| Single agent** | 1.5 | 120.2 | 1.3 | 101.5 | 0.34 | 127.7 |
| **GB-0669 + antiviral \| Combination<sup>a</sup>** | 0.042 | 119.1 | 0.072 | 103.9 | 0.44 | 122.4 |
| **Antiviral + GB-0669 \| Combination<sup>b</sup>** | 0.028 | 119.1 | 0.040 | 103.9 | 0.13 | 122.4 |

EC50 values were determined for each article tested as a single agent or in combination, defined as the concentration required to achieve 50% neutralization of infection. Data are representative of one experiment performed with three technical replicates. For combinations involving remdesivir, nirmatrelvir, or molnupiravir, EC50 values are representative of the GB-0669 dilution (combination<sup>a</sup>) or the corresponding antiviral compound dilution (combination<sup>b</sup>).
<sup>c</sup>Upper plateau represents the maximum % inhibition value derived from four-parameter logistic regression.

## Discussion

The results of this study demonstrate that the safety profile of GB-0669 in this healthy volunteer population was favorable and supports continued clinical development.

The safety profile observed in this study is reassuring and consistent with the known risks associated with the use of mAbs. One Grade 2 IRR that resolved with treatment was reported in the 1200 mg cohort and led to study drug discontinuation. Of note, IRRs are considered a well-known potential risk for mAbs as a class of therapeutics [21].

Antibody engineering involved an LS mutation in the Fc region to extend the half-life and promote translocation of the antibody to mucosal tissue, aimed at improving biodistribution in the lungs [15,16]. A similar design was used for the development of sotrovimab, a mAb used for COVID-19 treatment which remained efficacious until the emergence of the BA.2.86* variant of interest [22]. This enables single dosing, which is particularly advantageous for an approach that requires IV administration under clinical care with the cost, time, and patient burden involved.

*Ex vivo* measurement of serum live virus neutralization activity versus SARS-CoV-2 Omicron variants (BA.5.5 and XBB.1.5) supports advancement of GB-0669 studies into a 1200 mg dose, even under conservative assumptions. The neutralizing index model provides evidence that GB-0669 achieves therapeutic concentrations with one dose throughout the desired treatment interval. A key advantage of the neutralizing index method for calculating a therapeutic threshold is that the method links serum neutralizing titer directly to clinical efficacy with mAbs, without making any assumptions about tissue penetration [19].

Immunocompromised patients remain at high risk of COVID-19-related morbidity and mortality with few efficacious treatment options. This patient population often develops prolonged infections including a risk of persistent SARS-CoV-2 infection and the clinical complications associated with COVID-19 [23]. Persistent SARS-CoV-2 infection responds poorly to currently licensed therapies, with one case series reporting viral clearance in only 30% of immunodeficient patients following treatment with remdesivir monotherapy [24]. Consequently, an effective monoclonal-antibody therapy would constitute a critical therapeutic advance for immunosuppressed patients, providing a much-needed option to curb SARS-CoV-2 infection when endogenous immunity and existing antivirals prove insufficient. The clinical value of RBD-targeting monoclonal antibodies for COVID-19 treatment has already been demonstrated by therapeutic regimens such as bamlanivimab + etesevimab, casirivimab + imdevimab (REGEN-COV), sotrovimab and bebtelovimab, all of which reduced viral load and curtailed disease progression in immunocompromised patients whose vaccine-induced immunity was inadequate [25–30]. However, as RBD-targeting antibodies lost activity against emerging SARS-CoV-2 variants, their overall effectiveness declined [31]. Preclinical studies suggest that GB-0669 is unlikely to share this limitation and is expected to retain efficacy against newly emerging variants [14].

Combination therapy for SARS-CoV-2 infection in immunocompromised patients has been suggested as a strategy to prevent the development of therapeutic resistance [20]. This can involve the use of a mAb in combination with a small-molecule antiviral agent such as remdesivir [20]. Consistent with this approach, the data presented here demonstrate superior live virus neutralization when GB-0669 is used in combination with small-molecule antivirals.

### Limitations

In the post-pandemic era, neutralizing antibodies against SARS-CoV-2 in healthy volunteers are widely prevalent because of natural infection or vaccination, thus limiting the interpretation of *ex vivo* serum neutralization against the virus. We have attempted to address this limitation by reporting change from baseline for neutralizing titer, instead of reporting less meaningful absolute titers. There is therefore some uncertainty as to how these pharmacodynamic results may translate into a highly immunocompromised population with little or no serum titers against SARS-CoV-2.

## Conclusion

The totality of the safety, pharmacokinetic, and pharmacodynamic data support a single IV dose of 1200 mg GB-0669 for further investigation, including evaluation for treatment of COVID-19 in immunocompromised patients who are at high risk of COVID-19-related morbidity and mortality. The 1200 mg dose provides therapeutic threshold neutralizing activity across current variants of SARS-CoV-2. In addition, the data support the use of GB-0669 in combination with a small-molecule antiviral agent such as remdesivir, nirmatrelvir, or molnupiravir to further improve the neutralization profile. Such combinations have the potential to offer a new and durable treatment option to a vulnerable population with an unmet clinical need.

## Data Availability

All data produced in the present study are available upon reasonable request to the corresponding author.

## Acknowledgments

Melanie Colegrave, a consultant medical writer at Morula Health, provided writing and editorial support funded by Generate Biomedicines.

## Study approval statement

The Institutional Review Board (IRB) of Advarra gave ethical approval for this work.

