## Supplemental Material for "First-in-Human Study of a First-in-Class AI-Designed Monoclonal Antibody (GB-0669) Against the Conserved SARS-CoV-2 Spike S2 Stem Helix"

**Supplementary data**

***Pre-clinical toxicity study***

Forty cynomolgus macaques were randomly allocated into groups (3 males and 3 females per group, with an additional 2/sex/group for recovery) and received either a single (400 mg/kg) dose of GB-0669, single dose vehicle control, repeat (400/600 mg/kg) administration of GB-0669 (for a total of 2 doses given 3 weeks apart), or repeat administration of vehicle control. The following endpoints were evaluated: mortality, clinical signs, evaluation of skin reaction, body weights, body weight gains, ophthalmology, veterinary neurological examinations, respiratory rate, electrocardiographic examinations, clinical pathology parameters (hematology, coagulation, clinical chemistry, and urinalysis), organ weights, macroscopic and microscopic examinations including an expanded histopathologic evaluation and immunohistochemical (IHC) evaluation for GB-0669 levels in selected tissues (i.e., pituitary gland, lung, pancreas and prostate), and IHC evaluation for hormones in the pituitary gland (i.e., luteinizing hormone [LH] and thyroid stimulating hormone [TSH]).

*Pharmacokinetics*

GB-0669 was quantified in monkey serum using a validated ligand binding MSD assay. The target antigen, SARS-CoV-2 spike trimer, (Arco Biosystems, Newark, DE) was coated onto an MSD High Bind plate (Miso Scale Discovery, Rockville, MD). Minimum required dilution (MRD)-diluted standards, quality controls, blanks, and required samples were added to the plate, which was incubated and washed. Primary detection antibody, Goat anti-Human IgG polyclonal monkey adsorbed (Southern Biotech, Birmingham AL), was added, and the plate was incubated and washed. Secondary detection, Streptavidin-SulfoTag (Meso Scale Discovery, Rockville, MD) was added to the wells, and the plate was incubated and washed. Finally, a 2X MSD Read Buffer T was added to the wells. This assay resulted in a calibration range of 246 to 150,000 ng/mL.

***First-in-human study***

*Inclusion and exclusion of participants*

The inclusion criteria were as follows:

1. Must have given written informed consent before any study-related activities are carried out and must be able to understand the full nature and purpose of the trial, including possible risks and adverse effects.

2. Adult males and females, 18 to 55 years of age (inclusive) at screening.

3. Body mass index (BMI) ≥19.0 and ≤35.0 kg/m^2^.

4. Medically healthy without clinically significant abnormalities at screening and pre-dose on Day 1.

5. Negative SARS-CoV-2 rapid antigen or PCR tests prior to randomization.

6. Physical examination without any clinically relevant findings.

7. Systolic blood pressure in the range of 90 to 140 mmHg and diastolic blood pressure in the range of 50 to 90 mmHg after 5 minutes in seated, semi-recumbent, or supine position.

8. Heart rate (HR) in the range of 50 to 100 bpm after 5 minutes rest in seated, semi-recumbent, or supine position.

9. Body temperature, between 35.0°C and 37.5°C.

10. In the opinion of the investigator, no significant findings in serum chemistry, hematology, coagulation and urinalysis tests.

11. Female subjects of childbearing potential, defined as any woman who has experienced menarche and who is not permanently sterile or postmenopausal, must have negative blood pregnancy tests at screening (and negative urine pregnancy tests on Day 1) and must agree to use protocol-defined methods of contraception from screening through at least 90 days after study completion.

12. Male subjects must agree to use protocol-defined methods of contraception and agree to refrain from donating semen from screening through at least 90 days after study completion.

13. Have suitable venous access for intravenous infusion and blood sampling.

14. Be willing and able to comply with all study assessments and adhere to the protocol schedule and restrictions including staying overnight in the CRU.

The exclusion criteria were:

1. History or presence of significant cardiovascular, pulmonary, hepatic, renal, hematological, oncological, gastrointestinal, endocrine, immunologic, dermatologic, neurological or psychiatric disease, including any acute illness or surgery within the past 3 months determined by the principal investigator (PI) to be clinically relevant. Subjects with localized cancers treated with curative intent and not on active therapy are allowed.

2. Current infection that requires antibiotic, antifungal, antiparasitic or antiviral medications.

3. Acute illness or fever within 3 days before study enrollment (enrollment may be delayed for full recovery if acceptable to the investigator).

4. Abnormal EKG or any known cardiac condition determined by the PI to be clinically relevant.

5. Positive testing for any of the following: human immunodeficiency virus (HIV), hepatitis B surface antigen (HBsAg), hepatitis C antibodies (HCV).

6. Women currently pregnant, lactating, or planning a pregnancy between enrollment and final study visit, or intends to donate ova during such time period.

7. Male participants intend to donate sperm during this study or before the end of the study.

8. Any history of anaphylactic-type reaction to any substance or any history of infusion related reactions or any allergy to components of the study drug.

9. Blood donation or significant blood loss (i.e., > 500 mL) within 56 days prior to Day 1.

10. Plasma donation within 7 days prior to Day 1.

11. Use of more than an average of 5 packs/week of tobacco/nicotine-containing product within 6 months prior to Day 1. Subjects must agree to refrain from smoking for the duration of the study.

12. Excessive intake of alcohol, defined as an average daily intake of greater than 2 standard drinks for women and 4 standard drinks for men, (1 bottle of beer (375mL) is equivalent to approximately 1.4 standard drinks, 1 glass of spirits (30mL) is equivalent to approximately 1 standard drink and 1 glass (150mL) of wine is equivalent to approximately 1.5 standard drinks).

13. History of alcohol abuse, illicit drug use, physical dependence to any opioid, or any history of drug abuse or addiction within 2 years prior screening.

14. Individuals currently participating or planning to participate in a study that involves an experimental agent (vaccine, drug, biologic, device, or medication); or who have received an experimental agent within 1 month before enrollment of this study; or expect to receive another experimental agent during participation of this study.

15. Positive urine test for drugs of abuse or alcohol (legal amounts of tetrahydrocannabinol [THC] are allowed).

16. Unwilling or unlikely to comply with the requirements of this study.

17. Any condition or disease that, in the opinion of the Investigator, would pose a risk to participant safety or interfere with study evaluation, procedures or completion.

*Study Design and Subjects*

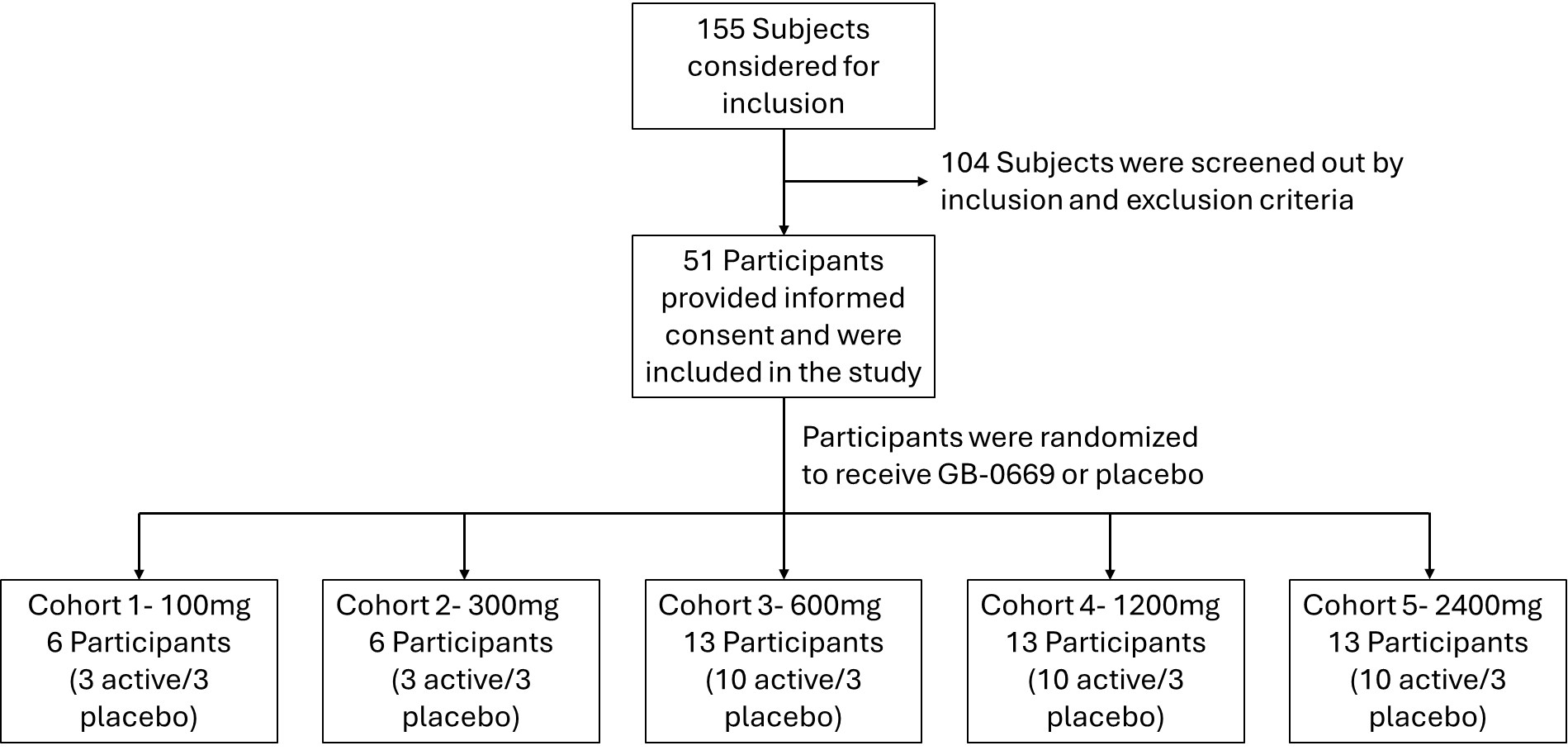
**Supplementary Figure 1.** Flow-chart showing the inclusion of participants in the first-in-human study.

**Supplementary Table 1.** Participants’ Demographics and Baseline Characteristics

|  | **GB-0669, n (%)** | | | | | **Placebo,**  **n (%)**  **N=15** |
| --- | --- | --- | --- | --- | --- | --- |
|  | **100 mg**  **N=3** | **300 mg**  **N=3** | **600 mg**  **N=10** | **1200 mg**  **N=10** | **2400 mg**  **N=10** |  |
| Age (years)  Mean (SD)  Median  Min-Max | 34.7 (10.50)  35.0  24, 45 | 38.0 (13.00)  38.0  25, 51 | 39.6 (10.35)  39.5  23, 53 | 35.8 (9.64)  34.0  23, 55 | 41.7 (8.97)  45.0  24, 52 | 33.2 (9.93)  31.0  23, 55 |
| Sex, n (%)  Female  Male | 2 (66.7)  1 (33.3) | 0  3 (100.0) | 6 (60.0)  4 (40.0) | 7 (70.0)  3 (30.0) | 5 (50.0)  5 (50.0) | 7 (46.7)  8 (53.3) |
| Race, n (%) ^1^  White  Black/African American  Asian  Multi-racial  Not reported | 1 (33.3)  1 (33.3)  1 (33.3)  0  0 | 2 (66.7)  1 (33.3)  0  0  0 | 7 (70.0)  2 (20.0)  0  1 (10.0)  0 | 3 (30.0)  5 (50.0)  0  1 (10.0)  1 (10.0) | 6 (60.0)  3 (30.0)  1 (10.0)  0  0 | 10 (66.7)  4 (26.7)  1 (6.7)  0  0 |
| Ethnicity, n (%) ^2^  Hispanic/Latino  Not Hispanic/Latino | 1 (33.3)  2 (66.7) | 2 (66.7)  1 (33.3) | 6 (60.0)  4 (40.0) | 3 (30.0)  7 (70.0) | 4 (40.0)  6 (60.0) | 6 (40.0)  9 (60.0) |
| BMI (kg/m^2^)  Mean (SD)  Median  Min, Max | 27.97 (4.65)  29.70  22.7, 31.5 | 27.43 (4.55)  26.10  23.7, 32.5 | 27.33 (3.19)  27.35  22.5, 32.1 | 26.66 (2.89)  26.30  22.2, 32.2 | 25.28 (3.66)  26.40  19.9, 30.7 | 28.41 (4.44)  29.70  20.1, 34.4 |
| BMI, body mass index; Max, maximum; Min, minimum; SD, standard deviation  ^1^No participants were reported as American Indian/Alaska Native, Native Hawaiian or Other Pacific Islander, or Other, so are not presented in this table.  ^2^No participants were listed under the categories of ‘Not reported’ or ‘Unknown’, so are not presented in this table.  *Safety*  Dose limiting toxicities (DLTs) were defined as any Grade 3 or greater AE or abnormal laboratory value assessed as related to the study drug in the view of the investigator that occurred during the study to Day 8. Severity grading was assessed using the Common Terminology Criteria for Adverse Events (CTCAE v5.0). If a DLT occurred, the iSRC reviewed all available safety data and recommended if dosing should continue or if the study was to be stopped. All participants were followed until the end of study (Day 302). The Safety Set consists of all participants treated with any dose of study drug. AEs by system order class/preferred term were coded according to the Medical Dictionary for Regulatory Activities (MedDRA, Version 26.0). | | | | | | |
